# Dynamic profile of severe or critical COVID-19 cases

**DOI:** 10.1101/2020.03.18.20038513

**Authors:** Yang Xu

## Abstract

In December 2019, a cluster of acute respiratory illness, now known as SARS-CoV-2 pneumonia, occurred in Wuhan, China. World Health Organization (WHO) declared the rapidly spreading coronavirus outbreak a pandemic on March 11, 2020, acknowledging what has seemed clear for some time — the virus will likely spread to all countries on the globe. As of February 11, 2020, the Chinese Center for Disease Control and Prevention (China CDC) has officially reported that there are 2.0% (889) asymptomatic cases, 2.3% (1,023) death cases, and 80.9% mild cases among 44,672 confirmed cases. 51.4% (22,981) were male and 48.6% (21,691) were female. Lymphopenia, in particular T lymphopenia, was common among patients with SARS-COV-2 in the observation. A notable drop in CD4 and CD8 lymphocyte counts occurred early in the course of the syndrome and was associated with adverse outcomes. The appearing a phenomenon of lymphocyte depletion (PLD) suggested severe adverse outcomes. The outcome observed: 60% had discharged and 20% had die.

## Introduction

In December 2019, a cluster of acute respiratory illness, now known as SARS-CoV-2 pneumonia, occurred in Wuhan, China.^1-5^ World Health Organization (WHO) declared the rapidly spreading coronavirus outbreak a pandemic on March 11, 2020, acknowledging what has seemed clear for some time — the virus will likely spread to all countries on the globe. As of February 11, 2020, the Chinese Center for Disease Control and Prevention (China CDC) has officially reported that there are 2.0% (889) asymptomatic cases, 2.3% (1,023) death cases, and 80.9% mild cases among 44,672 confirmed cases. 51.4% (22,981) were male and 48.6% (21,691) were female.^6^

In January, 2020, the 2019 novel coronavirus (SARS-CoV-2) was identified in samples of bronchoalveolar lavage fluid from a patient in Wuhan and was confirmed as the cause of the SARS-CoV-2 pneumonia.^1,2^ Full-genome sequencing and phylogenic analysis indicated that SARS-CoV-2 is a distinct clade from the betacoronaviruses associated with human severe acute respiratory syndrome (SARS) and Middle East respiratory syndrome (MERS). ^1,2^

COVID-19 has spread rapidly since it was first identified in Wuhan and has been shown to have a wide spectrum of severity. Recently, a report shows that SARS-CoV and SARS-CoV-2 shared the same functional host-cell receptor, angiotensin-converting enzyme 2 (ACE2).^7^ Furthermore, SARS-CoV-2 binds to ACE2 receptors in 10-20 fold higher affinity than SARS-CoV binds to the same receptors.^7^ Another report shows SARS-CoV-2 cell entry depends on ACE2 and TMPRSS2.^8^ Since ACE2 receptors on lung alveolar epithelial cells and enterocytes of the small intestine are dominant, lung alveolar epithelial cells or enterocytes of the small intestine may be an important susceptibility factor for humans.^9^

According to World Health Organization interim guidance on January 12, 2020, SARS-CoV-2 infection is classified as asymptomatic cases, mild and severe cases of pneumonia, and critical cases of pneumonia (ARDS, sepsis, septic shock). Severe cases of pneumonia are defined as patients with respiratory rate > 30 breaths/min, severe respiratory distress, or SpO_2_ < 90% on room air.^10^ Asymptomatic cases have been reported in China and Germany.^11,12^ Huang et al^3^ first reported 41 cases of SARS-CoV-2 pneumonia in which most patients had a history of exposure to Huanan Seafood Wholesale Market. Organ dysfunction (shock, acute respiratory distress syndrome [ARDS], acute cardiac injury, and acute kidney injury, etc.) and death can occur in severe or critical cases. Guan et al^4^ reported findings from 1099 cases of SARS-CoV-2 pneumonia and the results suggested that the SARS-CoV-2 infection clustered within groups of humans in close contact, and was more likely to affect older men with comorbidities. However, dynamic profile of COVID-19 infection and risk factors associated with COVID-19 pneumonia is not fully known.

## Methods

### Study Design and Participants

This retrospective observation study was done at affiliated hospitals of Shanghai University of Medicine & Health Sciences. This case series was approved by the institutional ethics board of Shanghai University of Medicine & Health Sciences (#2019-LCHZ-18-20190507). Written informed consent was waived due to the rapid emergence of this infectious disease. Identification of patients was achieved by reviewing and analyzing available electronic medical records and patient care resources. We retrospectively analyzed patients according to WHO interim guidance.^13^ Laboratory confirmation of SARS-CoV-2 infection was performed as previously described.^14^

### Real-Time Reverse Transcription Polymerase Chain Reaction Assay for SARS-CoV-2

A confirmed case of COVID-19 is defined as a positive result on high throughput sequencing or real-time reverse-transcriptase–polymerase-chain-reaction (RT-PCR) assay of pharyngeal swab specimens. Samples were collected for extracting SARS-CoV-2 RNA from patients suspected of having SARS-CoV-2 infection as described previously.^14^ In brief, the pharyngeal swabs were placed into a collection tube with 150 μL of virus preservation solution, and total RNA was extracted within 2 hours using the respiratory sample RNA isolation kit. Forty μL of cell lysates were transferred into a collection tube followed by vortex for 10 seconds. After standing at room temperature for 10 minutes, the collection tube was centrifugated at 1000 rpm/min for 5 minutes. The suspension was used for RT-PCR assay of SARS-CoV-2 RNA. Two target genes, including open reading frame 1ab (*ORF1ab*) and nucleocapsid protein (N), were simultaneously amplified and tested during the RT-PCR assay. Target 1 (*ORF1ab*): forward primer CCCTGTGGGTTTTACACTTAA; reverse primer ACGATTGTGCATCAGCTGA; and the probe 5′-VIC-CCGTCTGCGGTATGTGGAAAGGTTATGG-BHQ1-3′. Target 2 (N): forward primer GGGGAACTTCTCCTGCTAGAAT; reverse primer CAGACATTTTGCTCTCAAGCTG; and the probe 5′-FAM-TTGCTGCTGCTTGACAGATT-TAMRA-3′. The RT-PCR assay was performed using a SARS-CoV-2 nucleic acid detection kit. Reaction mixture contains 12 μL of reaction buffer, 4 μL of enzyme solution, 4 μL of probe primers solution, 3 μL of diethyl pyrocarbonate–treated water, and 2 μL of RNA template. The RT-PCR assay was performed under the following conditions: incubation at 50 °C for 15 minutes and 95 °C for 5 minutes, 40 cycles of denaturation at 94 °C for 15 seconds, and extending and collecting fluorescence signal at 55 °C for 45 seconds. A cycle threshold value (Ct-value) less than 37 was defined as a positive test result, and a Ct-value of 40 or more was defined as a negative test. These diagnostic criteria were based on the recommendation by the National Institute for Viral Disease Control and Prevention (China). A medium load, defined as a Ct-value of 37 to less than 40, required confirmation by retesting. Only RT-PCR confirmed cases were included in the analysis.

### COVID-19 IgG and IgM Assay

Goat anti-human IgG and IgM (62-8400, A18843) were purchased from ThermoFisher, Inc. COVID-19 recombinant antigen (R18850) was developed and purified at Meridian Life Science. The recombinant antigen (R18850) is receptor binding domain of SARS-CoV-2 Spike Protein. Several different designs of antigen were tested and optimized. Eventually, R18850 was picked into testing product. Bovine serum albumin (BSA), and goat anti-human IgG and IgM antibodies, rabbit IgG and goat anti-rabbit IgG antibodies, and 50 nm gold nanoparticle (AuNP) colloids (753645) were obtained from Sigma-Aldrich. NC membrane and plastic pad were obtained from IMS Medical Scientific, Inc (Zhejiang, China). The glass fiber conjugate (GFC) pad was obtained from Whatman, Inc. The phosphate buffer saline (PBS) was purchased from Sigma-Aldrich. Inactivated COVID-19 positive serum and negative serum samples of patients with were supplied by China CDC, China. To prepare the AuNP conjugate, SARS-CoV-2 recombinant protein dissolved in PBS (1 mg/mL) was added to the mixture of 1 mL AuNP colloid (50 nm in diameter, OD=1) and 0.1 mL of borate buffer (0.1 M, pH 8.5). After incubation for 30 min at room temperature, 0.1 mL of 10 mg/mL BSA in PBS was added to the solution to block the AuNP surface. After incubation for 15 min at room temperature, the mixture was centrifuged at 10,000 rpm and 4°C for 20 min. The supernatant was discarded, and 1 mL of 1 mg/mL BSA in PBS was added to the AuNP conjugate to be re-suspended. The centrifugation and suspension process were repeated twice, and the final suspension solution was PBS. The AuNP-rabbit IgG conjugates was prepared and purified by the same procedure. The Kit consists of 5 parts, including plastic backing, sample pad, conjugate pad, absorbent pad and NC membrane. Every component of the strip should be given a pretreatment described as follows: the NC membrane was attached to a plastic backing layer for cutting and handling. The goat anti-human IgM,IgG and goat anti-rabbit-IgG were immobilized at test M, G and control (C) line, respectively. Conjugate pad was sprayed with mixture of AuNP-COVID-19 recombinant antigen conjugate and AuNP-rabbit-IgG. Sample pad was pretreated with BSA (3%, w/v) and Tween-20 (0.5%, w/v) before use.

## Results

This is a continuous research observation of our previous patients with lymphopenia, included 6 male and 4 female patients.^14^ All patients received broad spectrum antibiotics. 6 patients received oseltamivir and lopinavir/ritonavir. Intravenous methylprednisolone at high dosage was used in patients with respiratory distress or progressive consolidations on their chest radiograph.

### Treatment

Of the 10 patients, 10 (100%) required oxygen support in the hospital (Table 1). Among 10 patients, all patients received empirical antibiotic treatment. Based on renal function at admission, 6 patients received oseltamivir and lopinavir/ritonavir treatment. Intravenous methylprednisolone at high dosage was used in patients with respiratory distress or progressive consolidations on their chest radiograph.

**Table 1.** Characteristics of severe or critical COVID-19 without comorbidities.

| Contents | All patients<br>(n=10) | Survivors<br>(n=8) | Non-survivors<br>(n=2) |
| --- | --- | --- | --- |
| **Laboratory characteristics** |  |  |  |
| White blood cell count, 10<sup>9</sup>/L | 4.3 (3.5-5.1) | 4.2 (3.2-5.0) | 4.6 (3.5-5.7) |
| Neutrophil count, 10<sup>9</sup>/L | 2.8 (1.9-4.5) | 2.6 (1.3-4.3) | 4.3 (2.0-4.8) |
| Lymphocyte count, 10<sup>9</sup>/L | 0.7 (0.6-0.9) | 0.8 (0.7-0.9) | 0.7 (0.6-0.8) |
| Hemoglobin, g/L | 138 (121-151) | 138 (126-150) | 137 (128-146) |
| Platelet count, 10<sup>9</sup>/L | 165 (135-219) | 167 (150-221) | 170 (121-219) |
| Prothrombin time, s | 12.3 (11.7-13.1) | 12.6 (11.7-13.4) | 18.1 (11.9-12.5) |
| Creatinine, μmol/L | 65 (49-69) | 59 (49-69) | 103 (95-110) |
| ALT*, U/L | 24 (20-39) | 21 (19-49) | 28 (21-35) |
| AST*, U/L | 26 (21-33) | 23 (17-35) | 30 (26-33) |
| Creatinine kinase, U/L | 94 (57-138) | 92 (57-134) | 120 (54-187) |
| Lactate dehydrogenase, U/L | 197 (164-289) | 199 (158-348) | 372 (174-571) |
| Hypersensitive troponin I, pg/mL | 3.3 (1.1-8.5) | 3.5 (0.7-5.6) | 4.1 (3.2-5.0) |
| D-dimer, mg/L | 0.5 (0.3-1.2) | 0.5 (0.3-0.9) | 2.4 (0.6-4.1) |
| Procalcitonin, ng/mL | 0.1 (0.1-0.1) | 0.1 (0.1-0.1) | 0.2 (0.1-0.3) |
| CRP, mg/L | 12.1 (3.6-29.1) | 10.1 (3.5-21.2) | 25.5 (21.9-30.1) |
| Interleukin-6, pg/mL | 17 (6-32) | 13 (9-33) | 21 (8-35) |
| **COVID-19 IgG and IgM Assay\*** |  |  |  |
| IgM | 4 (40.0%) | 3 (37.5%) | 1 (50.0%) |
| IgG | 3 (30.0%) | 2 (25.0%) | 1 (50.0%) |
| IgM + IgG | 9 (90.0%) | 7 (87.5%) | 2 (100%) |
| **Treatment** |  |  |  |
| Oxygen Support | 10 (100%) | 8 (100%) | 2 (100%) |
| Antiviral therapy | 6 (60.0%) | 6 (75.0%) | 0 (0.0%) |
| Antibiotic therapy | 10 (100%) | 8 (100%) | 2 (100%) |
| Corticosteroid therapy | 5 (50.0%) | 3 (37.5%) | 2 (100%) |
| **Clinical outcome** |  |  |  |
| In hospital | 2 (20.0%) | 2 (25.0%) | 0 (0.0%) |
| Discharge | 6 (60.0%) | 6 (75.0%) | 0 (0.0%) |
| Death | 2 (20.0%) | 0 (0.0%) | 2 (100%) |

\* COVID-19 IgG and IgM assay tested on day 15-30 of the observation; ALT=alanine aminotransferase; AST=aspartate aminotransferase.

### Lymphocytes

Of 10 patients, lymphopenia (absolute lymphocyte count < 1000/mm^3^) was noted in 10 (100% of patients) during their course of illness. Two patients were observed appearing a phenomenon of lymphocyte depletion (PLD, progressive lymphopenia), occurred in the day 5-10 of illness and reached its lowest point in the day 20-30. The lymphocyte count did not recover in the day 15-20 compared to other 8 patients, leading to the PLD till death occurred (Figure 1). Patient 1 (P1) had die in the day 32 of illness and patient 2 (P2) had die in the day 28 of illness.

**Figure 1.**
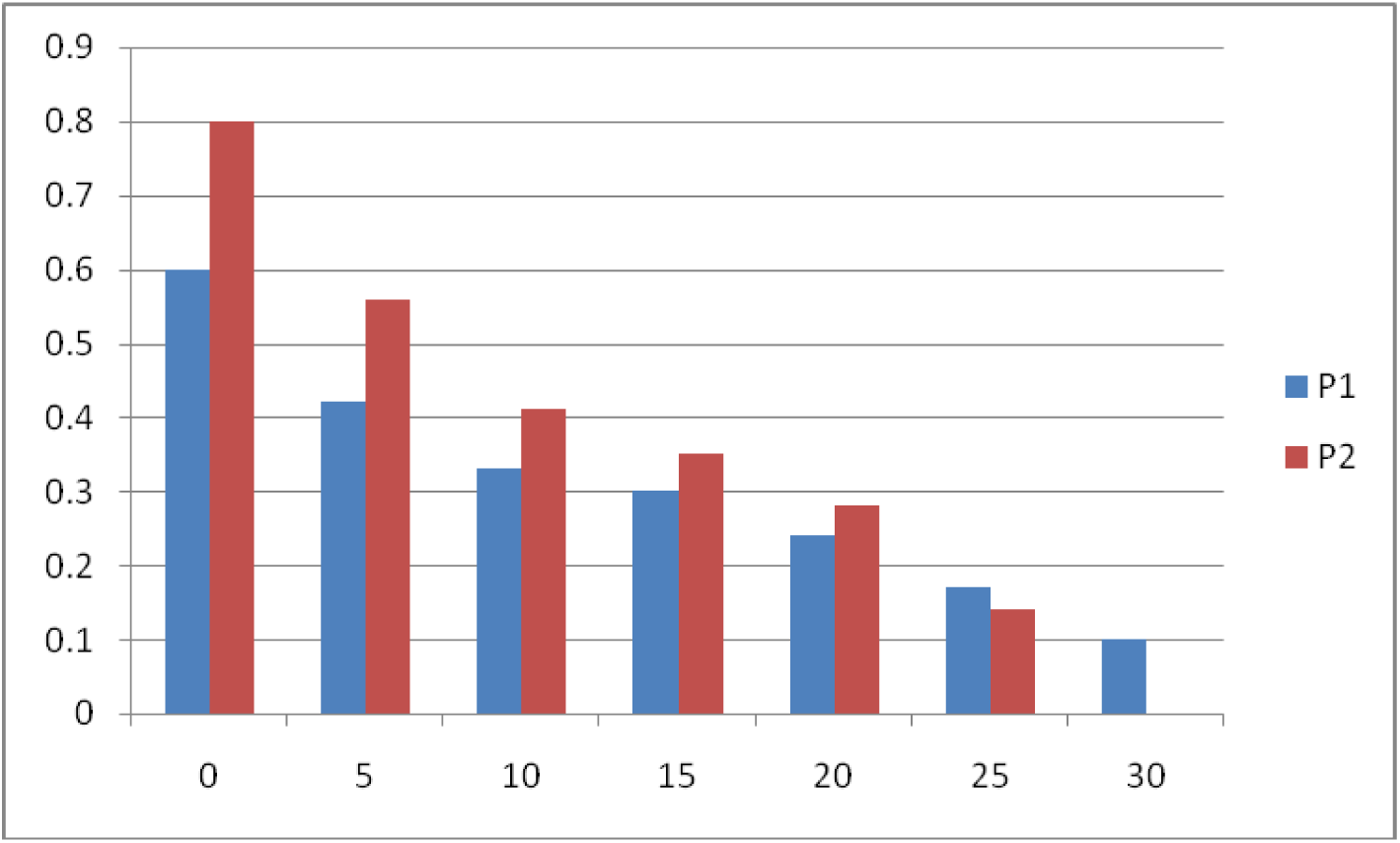
Proportion of patients with absolute lymphopenia during the course of SARS-COV-2. X-axis represents day and Y-axis represents cells 10^9^/L.

Subsets of peripheral blood lymphocytes in 10 patients were analyzed. Most patients had reduced CD4 and CD8 cell counts during the early phase of illness, which reached a trough on day 5 to 15 and recovered gradually afterwards (Figure 2, A). The ratio of CD4 to CD8 cells remained in the normal range.

**Figure 2.**
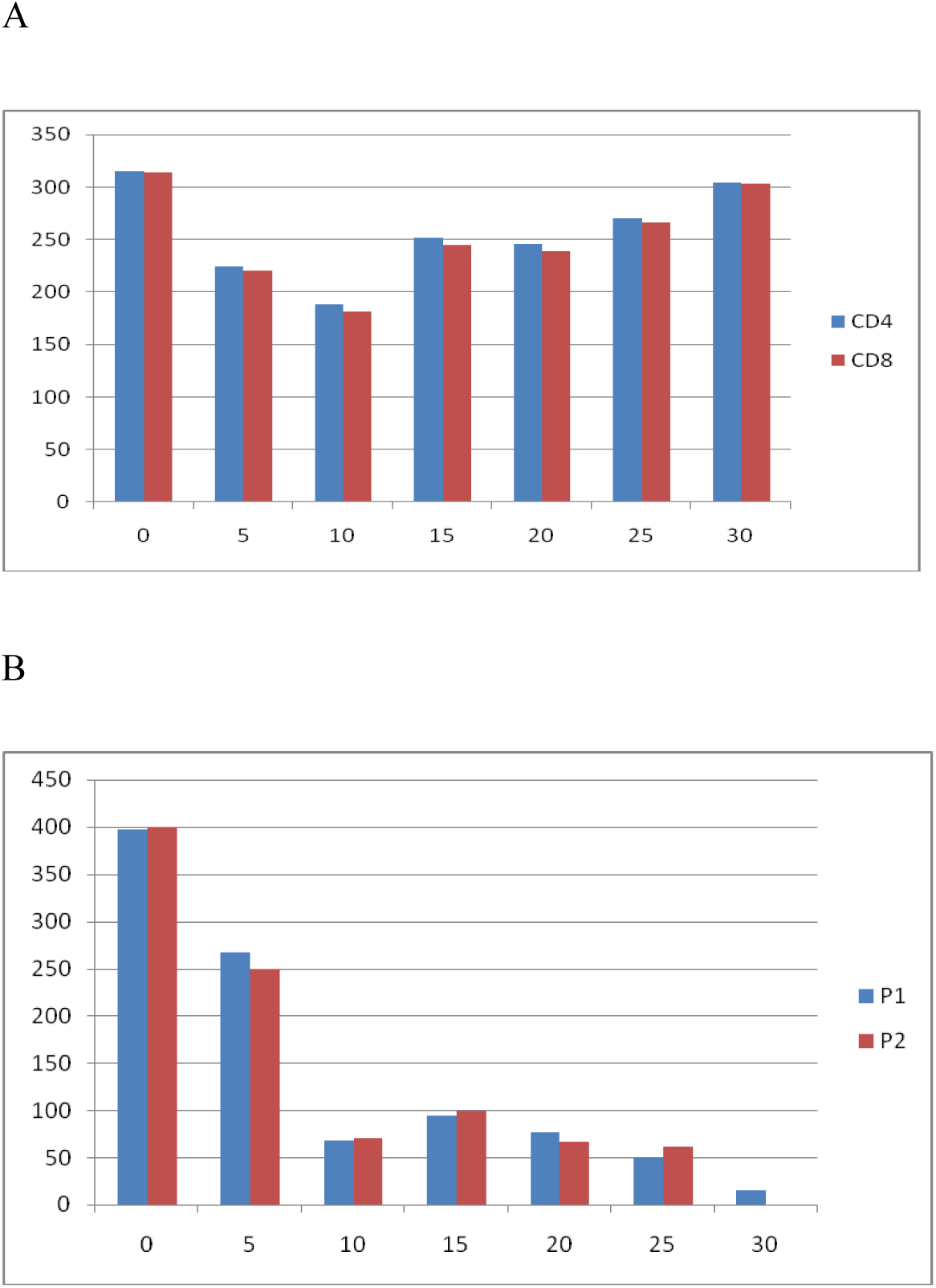
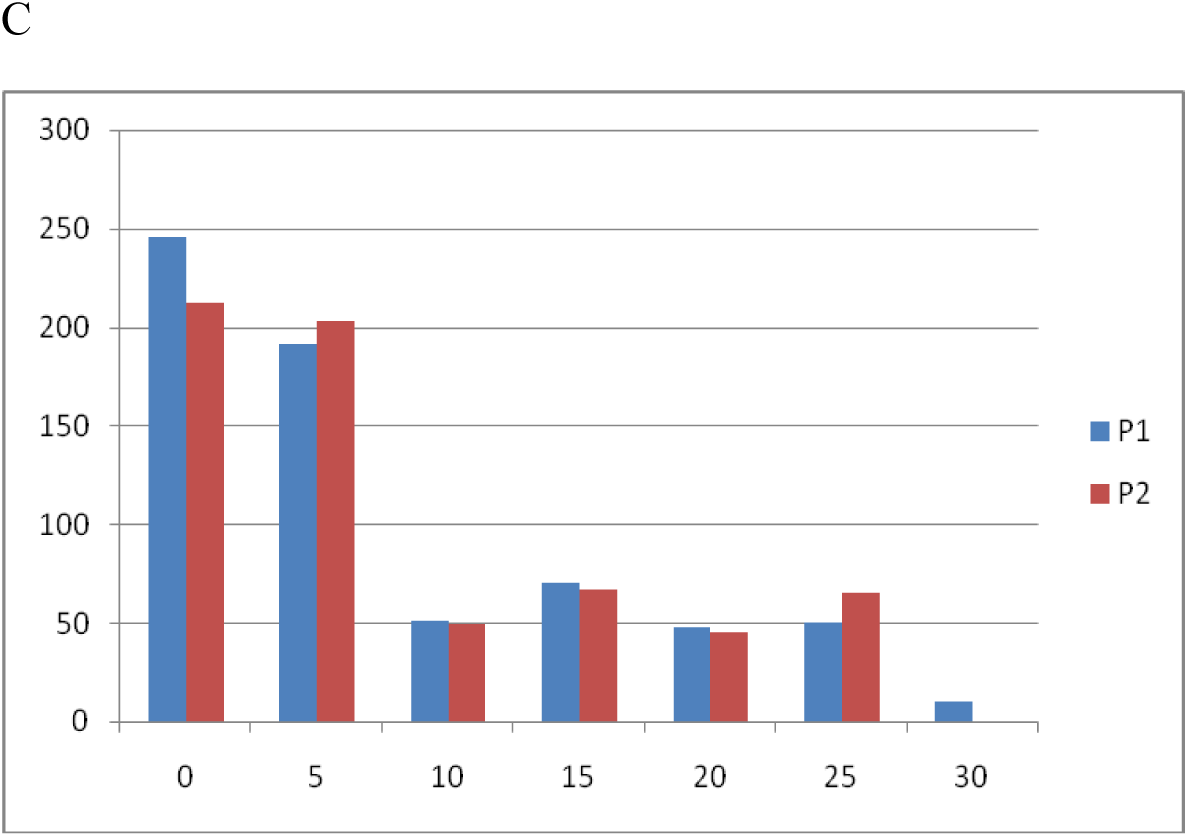
CD4 and CD8 lymphocyte subsets in peripheral blood against day of illness in 10 patients. X-axis represents day and Y-axis represents cells/mm^3^.

The CD4 and CD8 lymphocyte subsets of patient 1 (P1) and patient 2 (P2) did not recover in the day 15-20 compared to other 8 patients, leading to the PLD till death occurred (Figure 2, B, C).

## Discussion

Abnormal lymphocyte variables were common among patients with the new respiratory illness known as SARS-COV-2, which has caused a pandemic worldwide. Lymphopenia, the depletion of T lymphocyte subsets, and the PLD may be associated with disease severity linked to mortality. Studies of its effect on various body systems are crucial to the understanding of this disease.

Due to one transferred patient 7 in the study who was from other hospital co-infected with influenza A virus on day 9 of the observation, all other nine patients were tested for 9 additional respiratory pathogens. Patient 1-6 were already treated with oseltamivir and lopinavir/ritonavir at admission and all tested negative. Patient 8 was tested positive IgM for influenza A virus and oseltamivir and lopinavir/ritonavir treatment were added on day 15 of observation. Patient 1 and 2 were tested positive IgM and IgG for influenza A virus on day 25-30 of observation without treatment of antiviral therapy due to poor renal function.

The sensitivity of COVID-19 IgG and IgM assay tested on day 15-30 of the observation was 90% and 100% for non-survivors, suggesting that B cell function is still intact in non-survivors.

## Conclusions

Lymphopenia, in particular T lymphopenia, was common among patients with SARS-COV-2 in the observation. A notable drop in CD4 and CD8 lymphocyte counts occurred early in the course of the syndrome and was associated with adverse outcomes. The appearing a phenomenon of lymphocyte depletion (PLD) suggested severe adverse outcomes. The outcome observed: 60% had discharged and 20% had die. This study is the first observation of 40% patients co-infected with influenza A virus. Further studies to evaluate the mechanisms of these manifestations may help us to understand full pictures of this disease.

## Data Availability

we have all data referred to in the manuscript.

